# Olfactory loss is an early and reliable marker for COVID-19

**DOI:** 10.1101/2022.02.23.22271409

**Authors:** Behzad Iravani, Artin Arshamian, Johan N. Lundström

## Abstract

Detection of early and reliable symptoms is important in relation to limiting the spread of an infectious disease. For COVID-19, the most prevalent symptom is either losing or experiencing reduced olfactory functions. Anecdotal evidence suggests that olfactory dysfunction is also one of the earlier symptoms of COVID-19 but objective measures supporting this notion are currently missing. To determine whether olfactory dysfunction is an early sign of COVID-19, we assessed available longitudinal data from a web-based interface enabling individuals to test their sense of smell by rating the intensity of selected household odors. Individuals continuously used the interface to assess their olfactory functions and at each login, in addition to odor ratings, recorded their symptoms and result from potential COVID-19 test. A total of 205 COVID-19 positive individuals and 156 pseudo-randomly matched control individuals lacking positive test provided longitudinal data which enabled us to assess olfactory functions in relation to their test results date. We found that odor intensity ratings started to decline in the COVID-19 group as early as 6 days prior to test result date. Symptoms such as sore throat, aches, and runny nose appear around the same point in time; however, with a lower predictability of a COVID-19 diagnose. Our results suggest that olfactory dysfunction is an early symptom but does not appear before other related COVID-19 symptoms. Olfactory dysfunction is, however, more predictive of an COVID-19 diagnose than other early symptoms.

---

Olfactory dysfunction is a key symptom of the COVID-19 disease and symptom tracking studies have demonstrated that a sudden loss of olfactory functions is the most reliable symptom of the disease (Menni et al., 2020). Anecdotal evidence indicates that olfactory loss is an early symptom appearing before other symptoms but objective measures supporting this statement are currently missing.

Key to an individual’s attempt to limit the spread of any contagious disease is monitoring early disease symptoms. At the onset of the COVID-19 pandemic, fever and cough were reported as reliable early symptoms in non-hospitalized cases and considerable monitoring effort was globally focused on these two symptoms (Hu et al., 2020; Wei et al., 2020). However, olfactory dysfunction soon emerged as a symptom of interest (Tostmann et al., 2020) and we now know that nearly 50% of individuals with confirmed COVID-19 infection report complete loss of olfactory functions and an additional 10-20% report reduced olfactory sensitivity (Hannum et al., 2020; Gerkin et al., 2021). Given that nearly 70% of all individuals with COVID-19 lose either all or some olfactory functions at some stage of the disease, it is not surprising that a reduced sense of smell is the symptom with the highest odds ratio in non-hospitalized cases (Menni et al., 2020; Rudberg et al., 2020). Olfactory loss at some stage of the disease is so prevalent that loss of olfactory functions can be used to monitor increase of COVID-19 prevalence in a geographical area (Iravani et al., 2020; Pierron et al., 2020). The relationship between self-assessed and psychometrically assessed olfactory function is, however, poor (Landis et al., 2003). While most people will notice sudden and complete loss of olfactory function, awareness of a partial olfactory loss is still far lower than a perceptual loss in other sensory modalities, such as audition and vision. To reliably estimate olfactory loss, probing olfactory functions with actual odors is therefore needed.

At the onset of the pandemic, we provided the community with an online tool that enabled individuals to assess their olfactory performance using 5 selected common household odors from a list of 71 suggestions. Although the tool is anonymous to protect user privacy, individuals can continuously monitor their odor performance over time using a login mechanism. Importantly, at each login, the user completes a COVID-19 symptom check, reporting potential symptoms such as cough, fever, etc., as well as report any formal COVID-19 testing they had undergone as well as the outcome of the test. A number of individuals used the tool to continuously assess their olfactory functions and report their potential COVID-related symptoms, some of which contracted COVID-19 during their use of the tool, thereby creating a natural longitudinal experiment and data that enables direct comparisons between onset of olfactory dysfunction, COVID-related symptoms, and potential COVID-19 diagnose.

Utilizing this unique longitudinal sensory data, we assessed the hypotheses that a decline in olfactory functions occur before other COVID-related symptoms are reported by participants. Confirmation of this hypothesis would suggest that decline in olfactory function is not only an early symptom of COVID-19 but also a symptom that occurs before other common COVID-related symptoms.

## Materials and Methods

### Participants

A total of 5608 unique individuals enrolled, identified themselves as residing in Sweden, and entered complete data on the web-based data registry platform smelltracker.org during the 10 months between April–2020 to February–2021. We are here only assessing individuals from Sweden because our ethical permit for this assessment only covers Swedish residents and the time between COVID test and result distribution is uniform. Moreover, because we were only interested in assessing individuals who provided longitudinal odor data, we excluded all individuals who only completed one session, as well as 161 individuals who rated all odors consistently above 95 on a 0-100 scale, leaving a total of 1168 individuals. All the remaining individuals were above 18 years old, and their COVID-19 status were either confirmed with a PCR test, so-called C19+ (n = 205, 149 women and age: 43 ± 13) or not determined, so-called UC19. Given that the testing date distribution of UC19 cohort was different from that of the C19+, we pseudo-randomly selected individuals from UC19 (n = 152, 113 women and age: 45 ± 14) to comparably match the number individuals in two cohorts for a given month (**Figure 1A**). The study was approved by both the Israeli Edith Wolfson Medical Centre Helsinki Committee and the Swedish Institutional Review Board (Etikprövningsnämnden). Participants did not receive any form of monetary compensation for their participation and consent was waived. All aspects of the study complied with the Declaration of Helsinki for Medical Research involving Human Subjects.

**Figure 1.**
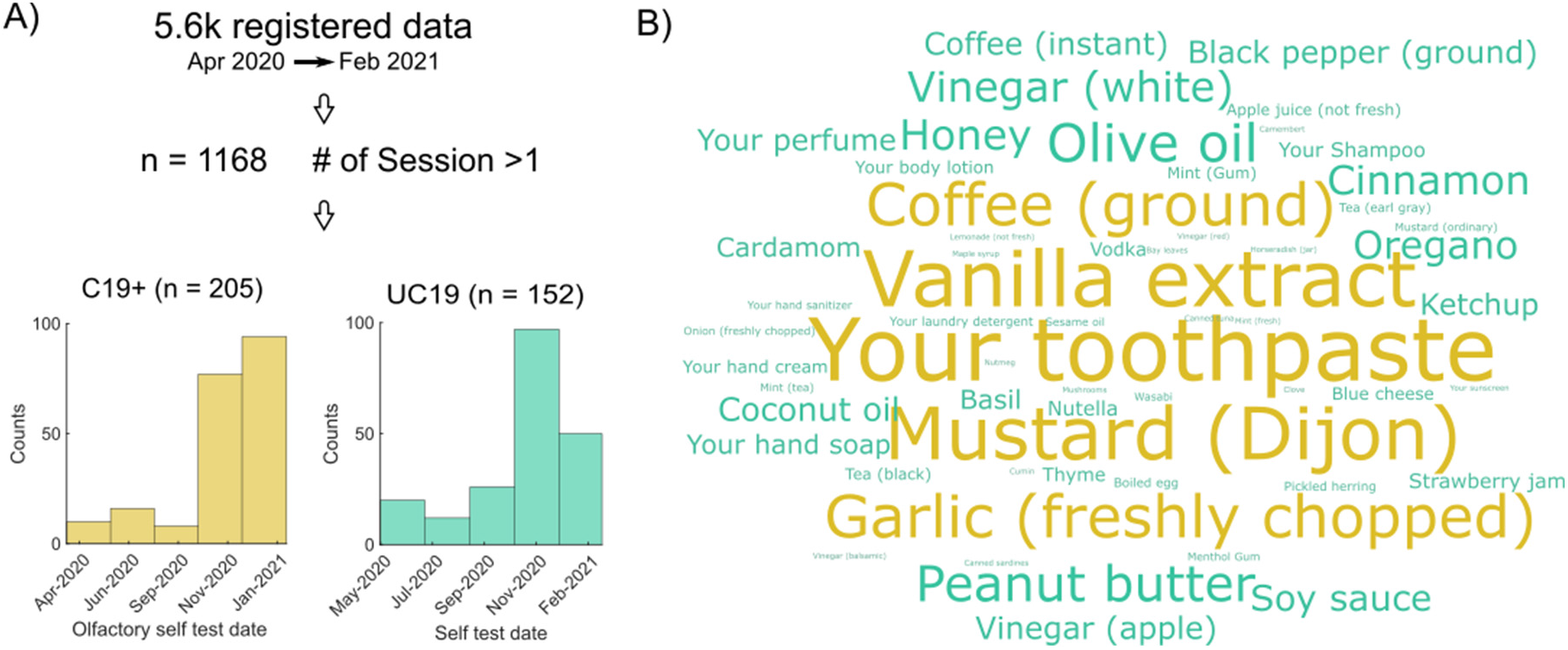
Data, self-test date density, and odors. **A**) Total number of individuals and the date range of data registration from which we included only individuals with more than one session of data registration. We pseudo-randomly sampled from unidentified COVID-19 status (UC19) cohort to match the population distribution of positive COVID-19 status (C19+) cohort. **B**) The word cloud shows the odor names that are rated and highlights the more frequently rated ones in a bigger and yellow font.

### Procedure and data collection

All the data collection was carried out via the Swedish version of the web-platform by which participants were able to create account to provide details regarding age, sex (Woman/Man/Other), and their COVID-19 test status (i.e., not tested, tested negative, tested positive). Particularly, regarding the COVID-19 status, If the participant provided no answer or marked “not tested”, we labeled them as “undermined”. Of note, we did not include participants who marked “tested negative” in the analysis to remove bias from our results due to the notion that these individuals got tested by experiencing symptoms that were not COVID-19 related. For repeated measurement, the web-platform allows individuals to repeatedly report their COVID-19 test status as well as self-test their odor performance. Specifically, for the odor performance test, participants chose 5 household odors from a list of 71 common household items. We had participants rate 5 odors to strike a balance between increased reliability, where more assessments render more reliable data (Kern et al., 2015), and low burden for participants to facilitate broad participation. The most frequently selected odors are illustrated in **Figure 2Figure 1B**. At repeated testing, the same 5 odors, freshly prepared, were used. Participants then proceeded to smell each odor and, on a separate page for each odor, rated their perceived intensity and pleasantness on visual analog scales, ranging from very weak/very unpleasant to very strong/very pleasant, respectively. These scales were coded as ranging from 0 (min) to 100 (max). Participants could smell the odors as often as they liked and there was no time pressure applied. We are here only focusing on odor intensity ratings. Moreover, in each session, participants were asked to report any experienced COVID-19 symptoms from a list of symptoms containing the following options: No symptoms, Fever, Cough, Shortness of breath or difficulty breathing, Tiredness, Aches, Runny nose, and Sore throat.

**Figure 2.**
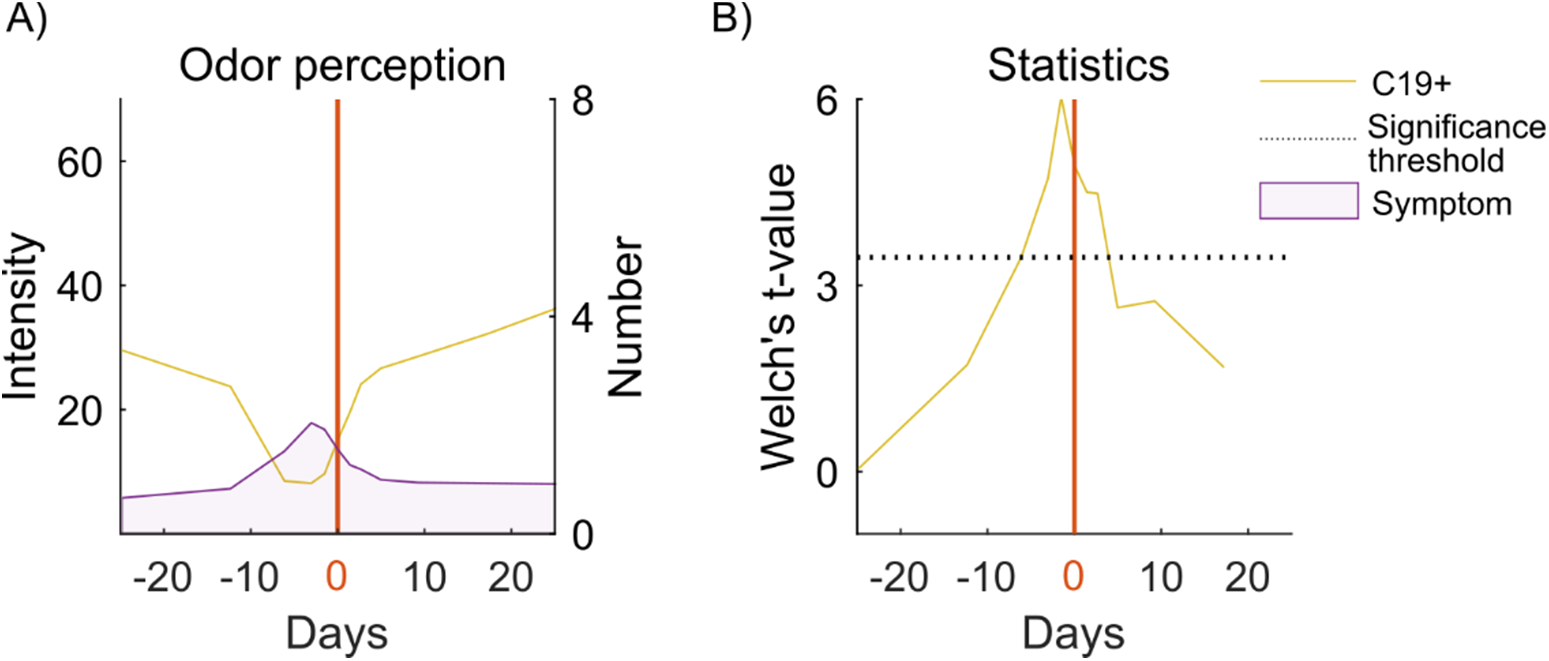
Odor intensity impairment and COVID-19. **A**) The yellow line indicates the median odor intensity rating as a function of days in relation to test result day (i.e., 0 and denoted with solid red line). Likewise, the purple area together with the right axis show aggregated number of symptoms, as a function of days, locked to test result day. **B**) The yellow line indicates the Welch’s t-value for the odor intensity measure as a function of days in relation to test result day. The significance threshold (p<.05) is shown with dotted black line.

### Data reduction and statistical analysis

We analyzed the C19+ odor intensity ratings to determine the time-course of the potential odor intensity impairment with respect to COVID-19 test result date. The interval during which the odor ratings were evaluated included a range between -25 to +25 days with the date of reported COVID-19 test result as day 0. This interval was logarithmically segmented to 13 bins and the ratings entered during each bin were averaged. Naturally, the number of individuals for each bins varies depending on the availability of the data for that specific bin with the maximum number individuals occurred in the bin that includes the test result date (i.e., equal to 205, the total number individuals in C19+). However, the number of individuals providing ratings in each bin varies between 4 to 205. To correct for the unbalanced distribution, we used Welch’s t-test wherein the inequality of variances is not a concern. Moreover, we created normative baseline values of intensity within the C19+ cohort by averaging ratings 60 days before or after the test result.

Identical data reduction was applied for assessing each of COVID-19 symptoms’ time-course as a function of days with respect to the test result date. However, the interval for this assessment was reduced to -10 to +10 days, using the date of reported COVID-19 test result as day 0, to achieve comparable statistical power. To assess differences between symptoms, we first estimated the null occurrence probability of each symptom in the UC19 cohort. Next, using a two-sided binomial test, we determined the corresponding z-value for each bin within the C19+ cohort. Significant and high z-values, for each day, indicate that the prevalence of this specific symptom is exclusive to COVID-19 whereas significant and low z-values denote that this specific symptom is not exclusive to COVID-19. Finally, we followed up on this analysis using logistic regression to assess the earliest day that each specific COVID-19 symptom was manifest in relation to test result date and if that given symptom was able to dissociate C19 from UC19. For each COVID-19 symptom, including odor intensity ratings, we fitted a logistic regression and compared the sensitivity, specificity, and the balanced accuracy, which is defined as the average of sensitivity and specificity. Nineteen unique individuals who were diagnosed with COVID-19 fulfilled the criteria for this analysis with enough longitudinal data. Consequently, we picked 21 random individuals from the UC19 cohort who registered data around the same day from a hypothetical test results day, here determined as the median of the reported test result dates (i.e., December 5^th^, 2020). Next, for each COVID-19 symptom, including odor intensity ratings, we used the fitted logistic regression model and determined the confusion matrix as well as the balanced accuracy for predicting C19+.

## Results

### Onset of reduced odor intensity perception might occur before positive COVID-19 test

We first sought to know whether measures of odor intensity had decreased before the individual underwent test for COVID-19. At the time of study, result after returned PCR test arrived within 2 days. To this end, we assessed the intensity ratings for C19+ across 25 days before and after the COVID-19 test result day to determine the curve of odor intensity impairment in COVID-19 over that extended time. Specifically, ratings of the 5 odors were averaged and time-locked to COVID-19 test result day. We found that the median of the odor intensity ratings started to decline in the C19+ group as early as 6 days prior to the test result date (i.e., denoted by 0 in **Figure 2A, 2B)** compared to C19+ baseline (**Figure 2B**), *t*(28.80) = 3.45, *p* < .01, *CI* = [12.43, 48.61].

### Onset of odor intensity impairment aligns with the earliest COVID-19 symptoms

Odor intensity ratings declined as early as 6 days prior to reported test result, thereby suggesting that a decline in odor intensity perception is an early sign of COVID-19. We therefore assessed whether odor intensity values were aligned with other COVID-19 symptoms. There was a significant negative association, *r*(14) = -0.95, *p* < .001, between median odor intensity ratings and number of COVID-19 symptoms for C19+ over time, as determined by a Spearman rank correlation. This finding demonstrates that odor intensity impairment aligned with COVID-19 symptom progression. Next, we asked whether the onset of decline in odor intensity perception occurred earlier than other non-odor related COVID-19 symptoms. To test this hypothesis, we first determined whether a COVID-19 symptom is significantly discernible on a specific day in the course of the disease by estimating the probability of reporting a specific symptom in the UC19 cohort. We found that probabilities of reporting COVID-19 symptoms in the UC19 group were as following (in descending order): Runny nose, probability [prob] = .32; Tiredness, prob = .31; Cough, prob = .21; Aches, prob = .12; Sore Throat, prob = .11; Fever, prob = .06; Shortness of Breath or Difficulty Breathing, p = .04. We considered these probabilities as our null hypothesis. Next, we assessed when, across the time course of ratings, each COVID-19 symptom in the C19+ group significantly stood out from the baseline (i.e., the null probabilities derived from the UC19 cohort) as a function of days locked to test result date. In other words, when each symptom might serve as an indication of COVID-19. We assessed this using two-way binomial tests separately for each symptom (**Figure 3**). We found that, in addition to olfactory intensity impairment that start to differentiate the groups 6 days prior to the test result date, Sore throats, (*z* = 4.25, *p* < .01), Aches (*z* = 3.30, *p* < .01), and Runny nose (*z* = 2.27, *p* < .03), are the earlies symptoms. It is worth mentioning that although the effect size for Runny nose is smaller than most of the aforementioned symptoms, it consistently stays above significance level for 3-4 days (Day -3: *z* = 3.12, *p* < .01). One other significant COVID-19 symptom that surpassed significance level was Fever (*z* = 2.15, *p* < .05) but at a slightly later time point compared to other symptoms, around -3 days in respect to test result day (**Figure 3**). It is worth noting that Shortness of breath or difficulty breathing did appear to increase earlier than -6 days, yet due to low number of observations at the early sessions, we were not able to statistically test the symptoms probability for a wider range of days.

**Figure 3.**
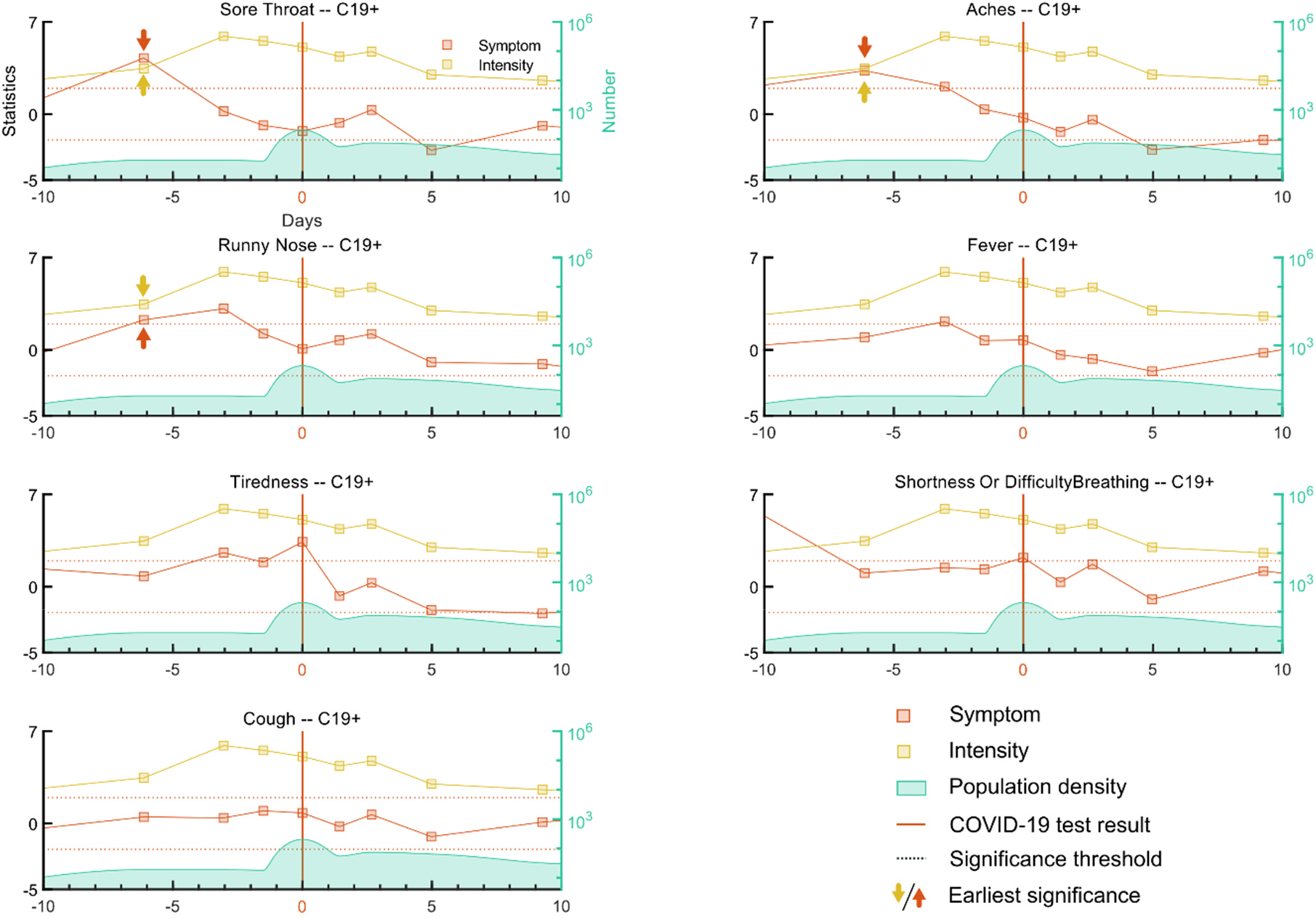
COVID-19 symptoms compared to intensity impairment time-courses. The time-course of 6 major COVID-19 symptoms including: Sore throat, Aches, Runny nose, Fever, Tiredness, Shortness of breath or difficulty breathing, and Cough time-courses, denoted by filled red connected squares were assessed and compared to the time-course of olfactory intensity impairment, denoted by filled yellow connected squares. The vertical red line at 0 depicts the test result day. The green distribution together with the green axis on the right side of the plots show the number of individuals for each specific day. Two dotted horizontal black lines show the significance threshold level. Red and yellow arrows show the earliest significant day for symptom and odor intensity impairment respectively.

Finally, we sought to determine which symptom in our data best predicted a COVID-19 diagnose on the -6 day using logistic regression models fitted to the data of each symptom across two cohorts. The confusion matrix for each symptom’s logistic model was computed to estimate sensitivity and specificity of that symptom (**Figure 4A**). We found that odor intensity impairment has the highest balanced accuracy of 70% followed by Runny nose with a balanced accuracy of 69%. Using Chi-squared test, we further found that odor intensity impairment, *χ*^2^ 13.1, *p* < .01, Runny nose, *χ*^2^ 6.61, *p* < .01, Aches, *χ*^2^ 5.91, *p* < .02, and Tiredness *χ*^2^ 5.06, *p* < .03, logistic models significantly outperformed the constant null model. The logistic model for Sore throat performed marginally better, *χ*^2^ 2.75, *p* < .10, then the constant null model. Other symptoms’ logistic models (all *p* >.34) were not significantly different from **Figure 4B**).

**Figure 4.**
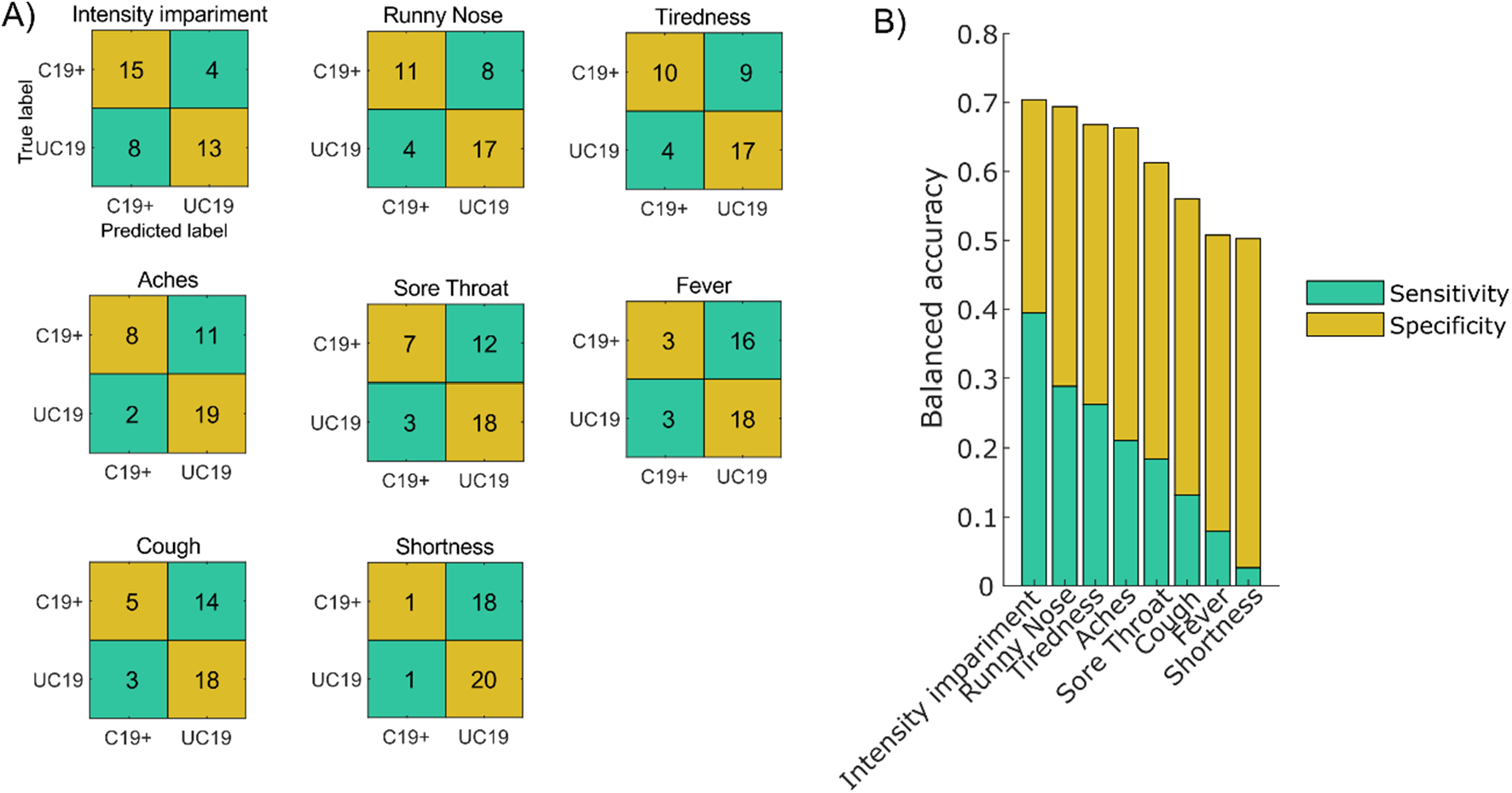
Logistic regression model of COVID-19 symptoms. **A**) Confusion matrices for the different COVID-19 symptoms. The vertical axis is the true label whereas the horizontal axis is the predicted label by the logistic model. The confusion matrixes are sorted according to their performance. **B**) Bars show the balanced accuracy, estimated as the average of sensitivity and specificity of each model. The lower and the upper segments of each bar represent the sensitivity and specificity respectively that contribute to the balanced accuracy.

## Discussion

We can here demonstrate that although reduced olfactory abilities are an early sign of COVID-19, it is not appearing earlier in the disease progression than several other symptoms of COVID-19. However, olfactory dysfunction was a symptom that demonstrated the highest predictability of COVID-19; a finding that has been demonstrated in several other studies using subjective measures.

Olfactory ratings started to clearly decline 6 days before participants indicated a positive test result. It is worth highlighting that all participants were regular data providers before their positive test result meaning that we were in a unique position to assess odor ratings before a potential test result might bias their ratings. It is not possible to definitely know, however, at what point in time participants were infected because test results might be communicated with different delays and participants did not provide information how much time had elapsed from test result and first post-result data entry. Nonetheless, it is clear that olfactory dysfunction was not only a common covid symptom but also an early one. Although it was a more reliable symptom during the early occurring strains of the SARS-CoV-2 virus (Rudberg et al., 2020), olfactory loss did not, however, seem to occur earlier than other common signs of COVID-19.

The main entry receptor for the SARS-CoV-2 virus is thought to be the angiotensin converting enzyme 2 (ACE2) receptor which is expressed throughout the human respiratory system with high density in the nasal epithelium and especially in the supporting sustentacular cells (Fodoulian et al., 2020; Hou et al., 2020; Muus et al., 2021). It is therefore not surprising that reduced olfactory functions is an early sign of COVID-19, appearing already 6 days before participants reported their positive test. Given the logical assumption that all participants using the webpage and its service were familiar with reports of the link between olfactory loss and COVID-19 and therefore might be positive towards not delaying their next olfactory assessment after news of positive COVID-19 tests, together with the average incubation time of the SARS-CoV-2 virus being reported as 5 to 6 days (McAloon et al., 2020), the decline in olfactory sensitivity occurs within the first days after infection.

Because the time of testing of included participants stretches over almost a year, several strains (Variants Being Monitored; VBM) of the SARS-CoV-2 can be assumed to have infected included participants. It is not possible to know exactly what proportions of VBMs dominated in our sample and when but the Wild-type strain, the B.1.1.7 (Alpha), and to a lesser extent, the B1.351 (Beta), were the dominating VBMs in the general Swedish population at the time in question (Public Health Agency of Sweden, 2022). Whether olfactory loss is an early and reliable sign of COVID-19 infection also for the Delta and Omicron VBMs is, at the time of writing in early February 2022, still debated. Tentative data originating from the verbal track-and-trace program in the United Kingdom suggests, however, that fewer individuals report subjective olfactory dysfunction after infection with the Omicron variant (Vihta et al., 2022). That said, these subjective data are collected already 1 to 2 days after a positive test and it is not yet determined whether potential lower numbers are due to a delay in onset of olfactory dysfunction or whether reports that the Omicron variant, in contrast to previous VBMs, often causes a nasal discharge or congestion might affect these early results (Vihta et al., 2022).

In the present study, we assess olfactory functions using intensity ratings of common household odors. In most studies on COVID-19 influence on the olfactory system, olfactory function has either not been assessed, assessed using subjective self-reports, or assessed with cued olfactory identification performance. Self-reports are a poor measure of olfactory function (Landis et al., 2003). While most people do notice sudden and complete loss of olfactory function, awareness of a partial loss is far lower than comparable perceptual loss in other sensory modalities like audition and vision. Cued identification performance alone is a crude measure of olfactory function that is most suitable to detect anosmia given the use of strong odors, that difficulty level is partly decided by the similarity between the presented odors and the lures on the cue card, and its partial reliance on cognitive and language skills (Larsson et al., 2004; Hedner et al., 2010). Therefore, to reliably estimate olfactory loss that does not border on anosmia, it is beneficial to probe aspects of olfactory function that is linked to the individual’s sensitivity. Odor intensity estimates are linked to the individual’s odor detection threshold (Cain, 1969). However, of higher relevance here is degree of fluctuation over time and based partly on the same data, we previously estimated the test-retest reliability of online odor intensity measure as .66, a value nearly identical to another study assessing test-retest of odor intensities (Kern et al., 2015). Moreover, this value is in-line with common odor detection thresholds where reliability between 4 time points been reported in the range of .43 to .85 (Albrecht et al., 2008).

In conclusion, we can demonstrate that for individuals infected by the SARS-CoV-2 virus, odor intensity ratings start to decline as early as 6 days prior their reported test result. However, other symptoms of COVID-19, such as aches, shortness of breath, and sore throat appears around the same point in time. These non-olfactory related symptoms display a lower predictability of a COVID-19 diagnose. Our results demonstrate that olfactory dysfunction is an early symptom of COVID-19 but not a symptom that appears before other related COVID-19 symptoms.

## Data Availability

All data produced in the present study are available upon reasonable request to the authors

## Acknowledgement

Funding provided by the Knut and Alice Wallenberg Foundation (KAW 2018.0152), the Swedish Research Council (2021-06527), and a donation from Stiftelsen Bygg-Göta för Vetenskaplig forskning, all JNL. Thank you to the Weizmann Olfaction Research Group for developing and maintaining the SmellTracker website.

## References

Albrecht, J., Anzinger, A., Kopietz, R., Schöpf, V., Kleemann, A.M., Pollatos, O., and Wiesmann, M. 2008. Test-retest reliability of the olfactory detection threshold test of the Sniffin’ sticks. Chem Senses. 33:461–467.

Cain, W.S. 1969. Odor intensity: Differences in the exponent of the psychophysical function. Percept Psychophys. 6:349–354.

Fodoulian, L., Tuberosa, J., Rossier, D., Boillat, M., Kan, C., Pauli, V., Egervari, K., Lobrinus, J.A., Landis, B.N., Carleton, A., et al. 2020. SARS-CoV-2 Receptors and Entry Genes Are Expressed in the Human Olfactory Neuroepithelium and Brain. IScience. 23:101839.

Gerkin, R.C., Ohla, K., Veldhuizen, M.G., Joseph, P.V., Kelly, C.E., Bakke, A.J., Steele, K.E., Farruggia, M.C., Pellegrino, R., Pepino, M.Y., et al. 2021. Recent Smell Loss Is the Best Predictor of COVID-19 Among Individuals With Recent Respiratory Symptoms. Chem Senses.46.

Hannum, M.E., Ramirez, V.A., Lipson, S.J., Herriman, R.D., Toskala, A.K., Lin, C., Joseph, P.V., and Reed, D.R. 2020. Objective Sensory Testing Methods Reveal a Higher Prevalence of Olfactory Loss in COVID-19-Positive Patients Compared to Subjective Methods: A Systematic Review and Meta-Analysis. Chem Senses. 45:865–874.

Hedner, M., Larsson, M., Arnold, N., Zucco, G.M., and Hummel, T. 2010. Cognitive factors in odor detection, odor discrimination, and odor identification tasks. J Clin Exp Neuropsychol. 32:1062–1067.

Hou, Y.J., Okuda, K., Edwards, C.E., Martinez, D.R., Asakura, T., Dinnon, K.H., Kato, T., Lee, R.E., Yount, B.L., Mascenik, T.M., et al. 2020. SARS-CoV-2 Reverse Genetics Reveals a Variable Infection Gradient in the Respiratory Tract. Cell. 182:429-446.e14.

Hu, Z., Song, C., Xu, C., Jin, G., Chen, Y., Xu, X., Ma, H., Chen, W., Lin, Y., Zheng, Y., et al. 2020. Clinical Characteristics of 24 Asymptomatic Infections with COVID-19 Screened among Close Contacts in Nanjing, China. MedRxiv.

Iravani, B., Arshamian, A., Ravia, A., Mishor, E., Snitz, K., Shushan, S., Roth, Y., Perl, O., Honigstein, D., Weissgross, R., et al. 2020. Relationship between odor intensity estimates and COVID-19 prevalence prediction in a Swedish population. Chem Senses. 45:449–456.

Kern, D.W., Schumm, L.P., Wroblewski, K.E., Pinto, J.M., Hummel, T., and McClintock, M.K. 2015. Olfactory thresholds of the U.S. Population of home-dwelling older adults: development and validation of a short, reliable measure. PLoS ONE. 10:e0118589.

Landis, B.N., Hummel, T., Hugentobler, M., Giger, R., and Lacroix, J.S. 2003. Ratings of overall olfactory function. Chem Senses. 28:691–694.

Larsson, M., Nilsson, L.-G., Olofsson, J.K., and Nordin, S. 2004. Demographic and cognitive predictors of cued odor identification: evidence from a population-based study. Chem Senses. 29:547–554.

McAloon, C., Collins, Á., Hunt, K., Barber, A., Byrne, A.W., Butler, F., Casey, M., Griffin, J., Lane, E., McEvoy, D., et al. 2020. Incubation period of COVID-19: a rapid systematic review and meta-analysis of observational research. BMJ Open. 10:e039652.

Menni, C., Valdes, A.M., Freidin, M.B., Sudre, C.H., Nguyen, L.H., Drew, D.A., Ganesh, S., Varsavsky, T., Cardoso, M.J., El-Sayed Moustafa, J.S., et al. 2020. Real-time tracking of self-reported symptoms to predict potential COVID-19. Nat Med. 26:1037–1040.

Muus, C., Luecken, M.D., Eraslan, G., Sikkema, L., Waghray, A., Heimberg, G., Kobayashi, Y., Vaishnav, E.D., Subramanian, A., Smillie, C., et al. 2021. Single-cell meta-analysis of SARS-CoV-2 entry genes across tissues and demographics. Nat Med. 27:546–559.

Pierron, D., Pereda-Loth, V., Mantel, M., Moranges, M., Bignon, E., Alva, O., Kabous, J., Heiske, M., Pacalon, J., David, R., et al. 2020. Smell and taste changes are early indicators of the COVID-19 pandemic and political decision effectiveness. Nat Commun. 11:5152.

Public Health Agency of Sweden. 2022. SARS-CoV-2 virusvarianter av särskild betydelse.

Rudberg, A.-S., Havervall, S., Månberg, A., Jernbom Falk, A., Aguilera, K., Ng, H., Gabrielsson, L., Salomonsson, A.-C., Hanke, L., Murrell, B., et al. 2020. SARS-CoV-2 exposure, symptoms and seroprevalence in healthcare workers in Sweden. Nat Commun. 11:5064.

Tostmann, A., Bradley, J., Bousema, T., Yiek, W.-K., Holwerda, M., Bleeker-Rovers, C., Ten Oever, J., Meijer, C., Rahamat-Langendoen, J., Hopman, J., et al. 2020. Strong associations and moderate predictive value of early symptoms for SARS-CoV-2 test positivity among healthcare workers, the Netherlands, March 2020. Euro Surveill. 25.

Vihta, K.-D., Pouwels, K.B., Peto, T.E., Pritchard, E., House, T., Studley, R., Rourke, E., Cook, D., Diamond, I., Crook, D., et al. 2022. Omicron-associated changes in sars-cov-2 symptoms in the united kingdom. MedRxiv.

Wei, W.E., Li, Z., Chiew, C.J., Yong, S.E., Toh, M.P., and Lee, V.J. 2020. Presymptomatic Transmission of SARS-CoV-2 - Singapore, January 23-March 16, 2020. MMWR Morb Mortal Wkly Rep. 69:411–415.

